# International Corneal and Ocular Surface Disease Dataset for Electronic Health Records

**DOI:** 10.1101/2025.01.18.25320772

**Authors:** Darren S. J. Ting, Stephen B. Kaye, Saaeha Rauz, the International Corneal and Ocular Surface Disease (C&OSD) Dataset Development Working Group

**Author notes:** **Corresponding author:** Professor Saaeha Rauz **Address:** Academic Unit of Ophthalmology, Birmingham and Midland Eye Centre, Department of Inflammation and Ageing, College of Medicine and Health, University of Birmingham, Dudley Road, Birmingham B18 7QU, United Kingdom. **Email:**.

## Abstract

**Background/Aims:** To provide a comprehensive and internationally standardised Cornea and Ocular Surface Disease (C&OSD) dataset for use in electronic health records (EHRs).

**Methods:** This was an international consensus study conducted through roundtable discussions involving 35 international experts specialising in the field of C&OSD. The Royal College of Ophthalmologists dataset guidelines were used to articulate initial C&OSD data elements template by curating data elements from validated published datasets obtained through scientific literature searches, and accessing existing international patient clinical and reported outcome recording instruments and registries. These included data elements recommended by the Dry Eye Workshop II, International Meibomian Gland Dysfunction Workshop, Ocular Surface Disease Activity and Damage Indices, the Cicatrising Conjunctivitis Assessment Tool, Limbal Stem Cell Deficiency Clinical and Confocal Grading, Chronic Ocular Manifestations in Patients with Stevens–Johnson Syndrome, and the UK Transplant Registry. Data elements pooled into an independent operational data model.

**Results:** A comprehensive generic dataset (common to all ophthalmology datasets) and C&OSD specific dataset was developed. Within the C&OSD dataset, several gateway disease datasets, such as atopic or allergic eye diseases, meibomian gland dysfunction, cicatrising conjunctivitis, chemical injury, dry eye, limbal stem cell deficiency, microbial or infectious keratitis, corneal erosion syndrome, and keratoconus, were established to streamline data entry for clinical audit and research purposes.

**Conclusion:** A comprehensive C&OSD dataset is provided which can be used by both generalist and specialist ophthalmologists. Adoption of the full dataset by EHR providers will lead to better interoperability and patient care and facilitate international research collaboration.

## INTRODUCTION

The advent of electronic patient or health records (EHR) has significantly transformed the landscape of healthcare systems. Since the inception of EHR in 1970s and subsequent development in 1990s, EHR systems have increasingly been adopted over the past decade across all branches of medicine, including ophthalmology, to replace traditional paper-based patient records.^1,2^ As of November 2023, 90% of the National Health Service (NHS) trusts in the UK have embraced EHR systems within their healthcare services as part of the digital transformation strategy of NHS Long Term Plan.^3,4^

EHR offers a multitude of advantages over traditional paper-based patient records.^5^ In 2016, the Global Observatory for eHealth (by the World Health Organisation) published the third global survey on eHealth,^6^ that highlighted the benefits of EHR systems in improving the quality, accuracy, reliability, and timeliness of patient information at the point of care. EHR systems can also provide linkages to other hospital information systems such as laboratory, pathology, imaging, and pharmacy information systems, and allow effective and efficient sharing of clinical information among different healthcare providers. Such shared-care records are an essential component underlying interoperability. In addition, EHRs enable the seamless capture of millions of clinical data points, which in turn can facilitate big data and artificial intelligence research.^7,8^ The availability of these real-world “big data” allows for a comprehensive and timely examination of the epidemiology, clinical characteristics, management, outcomes, and prognostic factors of diseases. Making better use of data and digital technology is a key component of the NHS Long Term Plan.^4^

Several barriers affecting implementation of EHR systems in healthcare have been reported in the real-world settings.^6,9,10^ These include the lack of funding, capacity, and infrastructure, legal issues related to data privacy and security, and time spent on the EHR systems by the clinicians (which may reduce patient-clinician interaction time). Moreover, many of the EHR systems are introduced for largely clinical and administrative purposes with secondary extraction for outcome monitoring, audit and research usage. Such EHR systems are designed to support generic medical data collection, with few specifically designed for medical or surgical specialties and subspecialties.

Ophthalmology-specific EHR systems such as Medisoft/Medisight (Medisoft Ltd, Leeds, UK) and OpenEyes (Apperta Foundation, Sunderland, UK) have been implemented into several centres. The content and standardisation of these do not sufficiently meet ophthalmic sub-speciality requirements and the lack of integration of these ophthalmology-specific EHR with generic medical EHR systems (e.g. Epic, Oracle, etc.) compromises patient care in the setting of multisystemic disease and conceals eye healthcare management strategies from other specialists. This not only hinders seamless access and exchange of ophthalmic and non-ophthalmic information between the electronic systems but may also pose major clinical risks due to incomplete patient information and assessment. The main functional components of an EHR are a clinical data repository, decision support system, order entry system, patient portal, and reporting system. Within the generic system, there should be a dedicated ophthalmic module/system (for each ophthalmology sub-speciality) to unify data capture across all EHR systems in order to standardise cross-institutional data collection and auditing. Moreover, not all ophthalmology units have adopted eye-dedicated EHRs, and clinical documentation is frequently restricted to free text entry or ‘form filling’ in generic EHR platforms.

In the UK, the ophthalmic EHR systems have supported secondary data analyses through the Royal College of Ophthalmologists (RCOphth) National Ophthalmology Databases (NOD). These currently focus on audit and research areas related to cataract surgery and age-related macular degeneration.^11–13^ There are no current NODs for corneal and ocular surface disease.

Datasets for the NOD provide specification for data collections and for data analyses for the benefit of improving standards of eye healthcare. They refer to a set of defined variables representing clinical information about a patient with a given condition and the format they should take. The RCOphth is committed to standardisation of data content (variables with grading scales), data type and format that meets Core Information Standards (CIS), ready for electronic use i.e., regardless of locality and which EHR system is being used, the curated data should be identical enabling subsequent pooling and comparisons of data to facilitate outcome monitoring and delivery of excellence in patient care.^14^

The purpose of this study is to: (i) establish an internationally agreed complete data itemisation proposed standard required to care for a patient with corneal and ocular surface disease (C&OSD), (ii) identify a library of data archetypes of commonly used items (such as visual acuity or intraocular pressure) that could be harmonised across all ophthalmological sub-specialty datasets, and (iii) embed functionality to extract sub-datasets to perform audit or report clinical outcomes such as following corneal transplant or after administration of high cost commissioned drugs or hospitalisation. Ultimately, we will seek designation of an Information Standards Notice (ISN) to demonstrate the minimum regulatory standard for reporting C&OSD in EHR systems.

## MATERIALS AND METHODS

The development and standardisation of international core dataset for C&OSD, the “Minimum Viable Information Standard (MVIS)”, followed the Professional Record Standards Body (PRSB) core information standards guidance that defines “best practice-based evidence with widescale consultation and input from users and relevant stakeholders”.^15^ Ethical approval was not required for establishing a C&OSD dataset proposed standard. All aspects of the workflow were conducted in accordance with the tenets of declaration of Helsinki.

### C&OSD proposed dataset standard workflow

The C&OSD EHR standard development group (EHR-SDG) consisted of 12 self-selected consultant ophthalmologists [SR (Lead), SBK (co-Lead), BA, CSL, DSJT, MKO’G, MR, ND, NL, OB, SA, and TB] from the Bowman Club (the UK corneal consultants consortium) and an international leaders working group of corneal and ocular surface disease experts by invitation AA, AJ, BS, BM, CB, CS, DSJ, FA, GI, GG, HSO, JC, JH, MM, MZ, NS, PR, RF, RD, SK, SLW, UJ, VR). Four rounds of online roundtable discussion, each lasting around 1-2 hours, were held amongst the UK expert panel between 27/09/2022 and 24/06/2023, with international consultation and validation held between 29/01/2023 and 15/03/2023. This was followed by one further (and final) meeting among UK experts on 20/03/2023 to discuss any remaining issues and finalise the international datasets prior to ratification by the RCOphth Quality and Standards Committee (version 1.0 published 16/07/2023). In the UK, a submission to the PRSB for a designated ISN is planned and it is intended that for all other countries a similar approach to the relevant patient record regulatory body will be undertaken. The conduct of this international consensus study is summarised in **Figure 1**.

**Figure 1.**
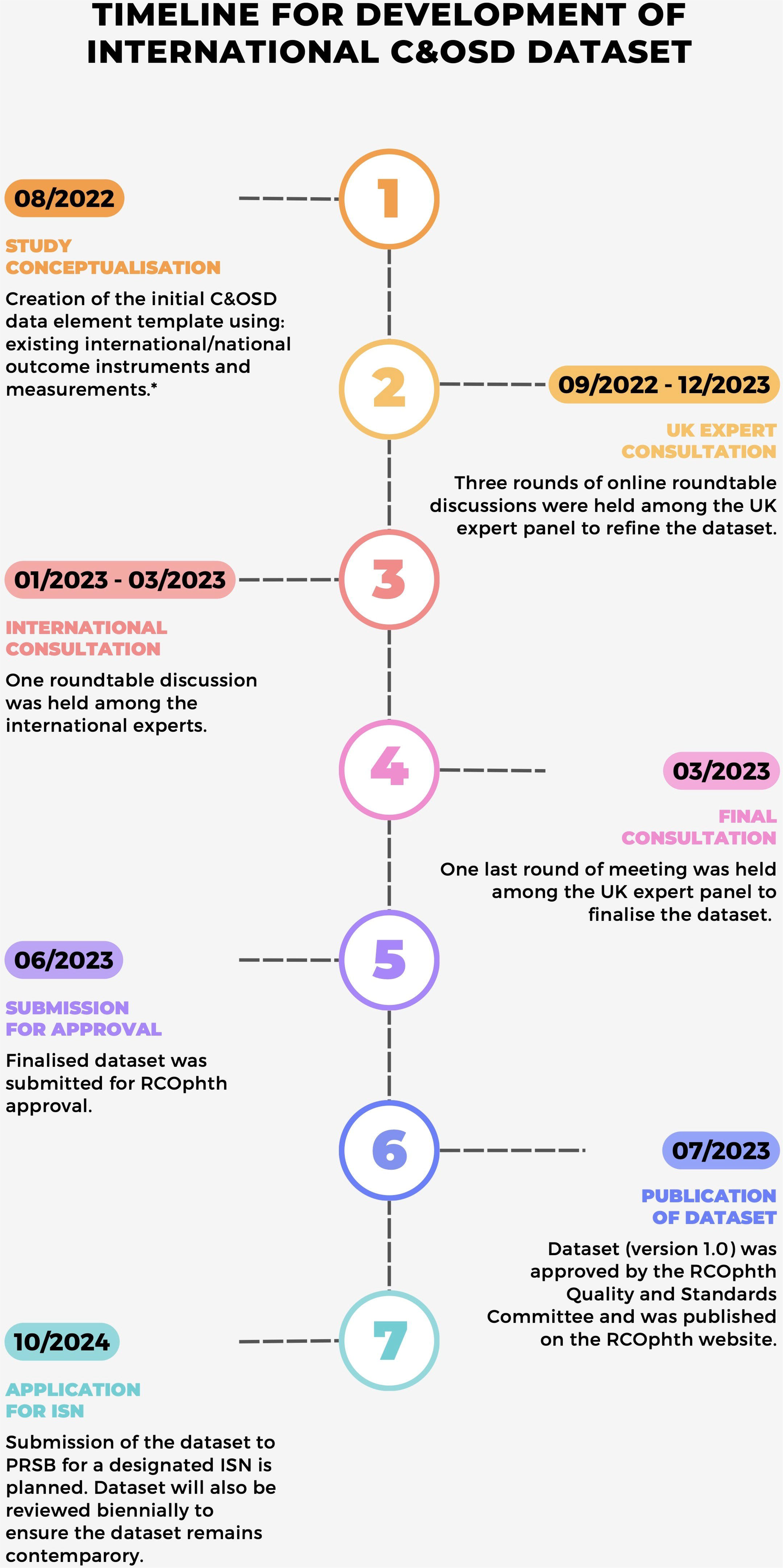
The timeline of the process of developing the international Corneal and Ocular Surface Disease (C&OSD) dataset. *These include Dry Eye Workshop II, International Meibomian Gland Dysfunction Workshop, Ocular Surface Disease Activity and Damage Indices, the cicatrising conjunctivitis assessment tool, Limbal Stem Cell Deficiency Clinical and Confocal Grading, Chronic Ocular Manifestations in Patients with Stevens–Johnson Syndrome, UK Transplant Registry – Ocular Tissue Outcome and Transplant Record, and RCOphth Cross-Linking Data Set. PRSB = Professional Record Standards Body; ISN = Information Standards Notice

### Construction and ordering of data elements

The RCOphth dataset guidelines from the Informatics Committee^14^ were used to articulate the initial C&OSD data elements template by curating data elements from validated published datasets for relevant conditions. The data elements were obtained through scientific literature search, or by access to existing international and national patient clinical and reported outcome recording instruments and registries.^16–23^ Validated datasets with health outcomes measurements or those agreed through large international consensus groups or the international consortium of health outcomes measurement (ICHOM) were included. Data elements for the assessment of corneal and ocular surface disease recommended by the Dry Eye Workshop II,^16^ International Meibomian Gland Dysfunction Workshop,^17^ Ocular Surface Disease Activity and Damage Indices,^18^ the cicatrising conjunctivitis assessment tool,^19^ Limbal Stem Cell Deficiency Clinical and Confocal Grading,^20^ Chronic Ocular Manifestations in Patients with Stevens–Johnson Syndrome,^21^ UK Transplant Registry – Ocular Tissue Outcome and Transplant Record^22^ (to substitute local transplant registry dataset), and RCOphth Cross-Linking Data Set^23^ (to substitute local CXL dataset) were extracted. Patient-reported outcome measures such as OSDI, IDEEL, and SANDE, were also included.

Data elements were pooled into an independent operational data model created with Microsoft Excel hosted on Sharepoint server. Duplicates were removed and a library of commonly used items (such as visual acuity or intraocular pressure) that could be harmonised across all ophthalmological sub-specialty datasets were identified and labelled as ‘generic’. Those specific to C&OSD were ordered into a coherent format using an explicit, empirically based approach to defining and naming relevant clinical constructs and assigning category and data type.^14^

Category assignment comprised of three categories: ‘Mandatory’ (data items which are essential for all applications, and must be collected), ‘Desirable’ (advised as valuable for audit or knowledge extraction purposes) and ‘Optional’ (data items which are required for some applications, and may be collected)

Data type assignment used structured query language (SQL) for managing and manipulating data in relational databases summarised below:

− TYPE: Description
− NULL: A special entity representing an uncertain or unassigned value
− INTEGER: An integer value, normally unsigned (i.e. zero or positive values only)
− FLOAT: A floating-point value, positive or negative (avoid spurious precision)
− BOOL: A value representing true or false
− STRING: A value containing text (alphanumeric data) of unspecified length
− ENUM: A value which represents one of a limited range of values
− DATE: A value representing a date
− DATETIME: A value representing a date and time
− LIST: An entity containing one or more values

Drawing templates were also adapted from the existing datasets or created new.

## RESULTS

A total of 35 international experts (from 15 countries) in the field of C&OSD were included in this study (**Supplementary Table 1).** Prior to each meeting, the dataset was distributed for comment and to identify missing, redundant or duplicated data fields, conditions or diseases that were not captured by the data fields, terminology (in particular if country specific), alignment with consensus standards for reporting and analysis of data (e.g., refractive and keratometric data) together with a navigational information video (**Supplementary Video 1**). The data fields were then amended with various elements being introduced at different stages and were further discussed at each meeting as an iterative process.

A full dataset was developed after five rounds of roundtable meetings (**Figure 2**). This dataset was then divided into generic dataset (which is common to all ophthalmology datasets) and C&OSD specific dataset. The generic datasets included four main domains, encompassing demographics, previously known general/ophthalmic diagnoses, general medical examination, and general ophthalmic examination. C&OSD included existing UK national datasets for corneal cross-linking, corneal transplant, and serum eye drops, as well as gateway diseases, which were brought in during the first national consultation. Nine gateway diseases (and their associated specific datasets) were created to facilitate specific data collection. These included blepharitis, dry eye disease, conjunctivitis (including simple, allergic/atopic, and cicatrising conjunctivitis), microbial/infectious/HSV keratitis, keratoconus, recurrent corneal erosion syndrome, chemical eye injury, and limbal stem cell deficiency (**Figure 2**). In order to account for colloquial preferences and to be inclusive for similar terms used for common conditions, subheadings included such additional colloquial terms. For example, corneal ‘ulcer’ was separated into (i) corneal abscess/infiltrate and (ii) corneal ulcer. Several drawing templates were created for facilitating the documentation of various C&OSD.

**Figure 2.**
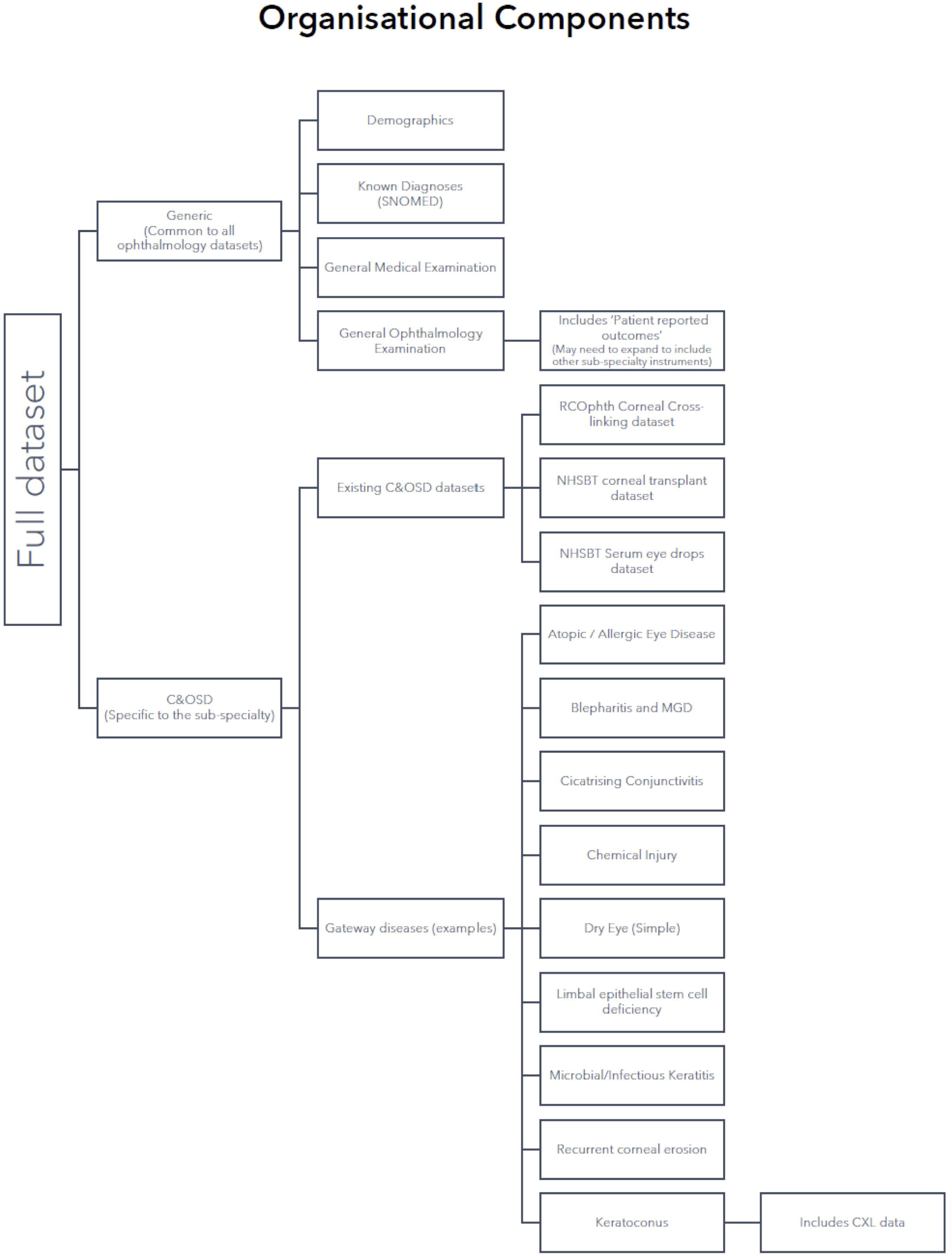
Organisational components of the full dataset, including examples of common putative gateway diseases and Corneal and Ocular Surface Disease (C&OSD) dataset.

Full details of the C&OSD datasets are accessible in the supplement and via the following link to The RCOphth Clinical Data Sets webpage (https://www.rcophth.ac.uk/standards-and-guidance/audit-and-data/clinical-data-sets/) where the dataset will be maintained and updated on a minimum of a biennial basis.

## DISCUSSION

Standardisation is the process of creating, issuing and implementing standards. A standard is a document, established by consensus and approved by a recognised body. It provides rules, guidelines or characteristics for activities or their results so that they can be repeated.^24^ The establishment of a minimum clinical dataset serves as a critical component of the healthcare system where it helps ensure the delivery of high quality care to individual patients as well as enable standardised clinical data collection, audit and research.^25^

This work represents the first international clinician-driven, standardised dataset for C&OSD. The proposed dataset is suitable for both general ophthalmologists and those with special interest in C&OSD. In addition, the involvement of experts from around the world ensured the comprehensiveness and generalisability of the dataset to be adopted in any country. The development of this seminal C&OSD dataset may not only help standardise the documentation of various C&OSD globally, but also enable interoperability and better patient care as well as facilitate collaborative multi-centre big data research on these conditions in the future.

To date, a few ophthalmic EHR systems have been developed and implemented across the world.^7^ One of the most significant examples is the Intelligent Research in Sight (IRIS) Registry, a US-based ophthalmic EHR registry established by the American Academy of Ophthalmology in 2014.^26,27^ As of October 2021, the IRIS Registry had captured data from >70 million patients (with >400 million patient visits) contributed by around 16,000 eye clinicians, serving as a powerful resource for benchmarking clinical performance and patient care/outcomes, epidemiological studies, scientific discovery, and big data research.^27^ In addition, the availability of high-quality big data provide invaluable opportunities for artificial intelligence (AI) research.^7,28^ So far, the IRIS Registry has enabled research in nearly all branches of ophthalmology, including corneal and ocular surface diseases.^29–33^ For instance, by analysing the data from >5 million patients in IRIS registry, Singh et al.^33^ were able to determine the case frequency of corneal opacity (6.5% of all patients) and identify the underlying causes and prognostic factors for poor visual outcomes, providing important insights into the epidemiology and unmet clinical/research needs of this important condition. Similarly, Das and Basu^34^ were able to identify >20,000 patients presented with epidemic keratoconjunctivitis and characterised the clinical features and outcomes using data of >2 million patients derived from EHR, improving the understanding of this prevalent disease. The Save Sight Dry Eye Registry, operational since 2020, has collected global real-world outcomes on the most common ocular surface disease (i.e. dry eye disease) and the Save Sight Keratoconus Registry on the outcomes of keratoconus since 2015.^35,36^ These registries have discovered that the impacts on quality of life of keratoconus are greater than macular disease, reported on disease natural history, crosslinking outcomes and treatment patterns in dry eye disease.^35,37–39^ The registries have also enabled clinicians to benchmark their practice and obtain credit towards continuing professional development accreditation.

Bearing in mind the demanding volume of C&OSD patients presenting to clinics and hospitals and the amount of time required for documentation using EHR systems,^10,40^ a number of gateway diseases were created to improve the clinical documentation of several prevalent and important C&OSD. In particular, we included common C&OSD conditions such as blepharitis, dry eye disease, conjunctivitis (including simple and allergic/atopic conjunctivitis), infectious (microbial and HSV) keratitis, keratoconus, recurrent corneal erosion syndrome, and chemical eye injury, as well as more specialised or rare conditions like cicatrising conjunctivitis and limbal stem cell deficiency. Other gateway diseases can be customised peculiar to the centre and clinical needs, e.g. Fuchs’ endothelial dystrophy, ocular surface neoplasia, and others. The Save Sight Keratoconus and Dry Eye Registries data fields are based on parsimony to ensure capability of data collection with everyday clinical practice.^41^ Overtime review of data entry using the C&OSD dataset based on parsimony may enable more efficient data entry.

One of the main intentions for the creation of the C&OSD dataset is to provide a dataset that is comprehensive and that can only work if it is adopted in full. Many of the EHRs that have been developed and supplied off the shelf have been selective in the data fields and/or have amended and modified the fields and terminology. This means that the components of many conditions are not adequately captured, and the lack of consistency severely limits interoperability and patient care. Provision of a common C&OSD dataset that is supported by international experts provides the EHR providers with a unique opportunity to improve patient care and facilitate knowledge exchange. While it is hoped that this C&OSD dataset is voluntarily adopted by EHR providers, it is also intended that the dataset becomes mandated. Each country will have their own mechanisms for this. In the UK, it will be achieved via an ISN. Alignment with Continuing Professional Development accreditation or outcome reporting for high cost or novel medical intervention requirements in other countries may facilitate uptake. The dataset that is provided, however, is not meant to be static and will need to be updated to keep abreast with advances in field and to include improvements with a set minimum biennial review cycle Similarly, it is essential that the views of new experts are included. In the future, the integration of AI and clinical registries may facilitate big data research using the C&OSD dataset.^7,28^ In order, to ensure the data remains up to data and utilises available technology, the dataset will be reviewed every 2 years under the governance of the RCOphth who will maintain the dataset and to whom comments and corrections can be submitted.

In conclusion, we have developed a comprehensive C&OSD dataset for both the generalists and specialists, through an international taskforce. We envisage that the adoption of the full dataset by EHR providers will lead to better interoperability and patient care and facilitate wider international research collaboration in the future.

## Data Availability

All data produced in the present work are contained in the manuscript.

## Notes

**Funding/support:** DSJT acknowledges the support from Birmingham Health Partners (BHP) Clinician Scientist Fellowship. SR is funded by the National Institute for Health and Care Research (NIHR) under its Invention for Innovation (i4i) (II-LA-1117-20001); UKRI Medical Research Council Experimental Medicine Award (MR/X019195/1). The views expressed are those of the author(s) and not necessarily those of the NIHR or the Department of Health and Social Care or MRC. JC acknowledges funding from the NIH grants (EY021558 and EY013124).

**Conflict of interest:** AA receives consultant fees and grant support from Santen, Inc., Japan. All other authors have no conflict of interest to declare.

### Competing Interest Statement

AA receives consultant fees and grant support from Santen, Inc., Japan. All other authors have no conflict of interest to declare.

### Funding Statement

DSJT acknowledges the support from Birmingham Health Partners (BHP) Clinician Scientist Fellowship. SR is funded by the National Institute for Health and Care Research (NIHR) under its Invention for Innovation (i4i) (II-LA-1117-20001); UKRI Medical Research Council Experimental Medicine Award (MR/X019195/1). The views expressed are those of the author(s) and not necessarily those of the NIHR or the Department of Health and Social Care or MRC. JC acknowledges funding from the NIH grants (EY021558 and EY013124).

