## Supplementary Table 1 for "International Corneal and Ocular Surface Disease Dataset for Electronic Health Records"

**Supplementary Table 1.** Summary of countries of the international experts involved.

Countries

Australia

Austria

Canada

Denmark

France

Germany

India

Italy

Japan

New Zealand

Pakistan

Russia

Singapore

United Kingdom

United States of America
