## Supplementary Table 2 for "International Corneal and Ocular Surface Disease Dataset for Electronic Health Records"

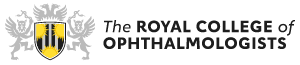

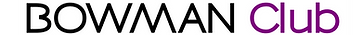

**Cornea and Ocular Surface Disease**

**Datasets for Electronic Patient Records**

**Cornea and Ocular Dataset**

**Please double click on the icon:**

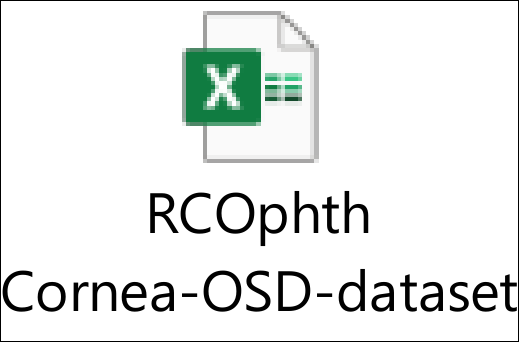

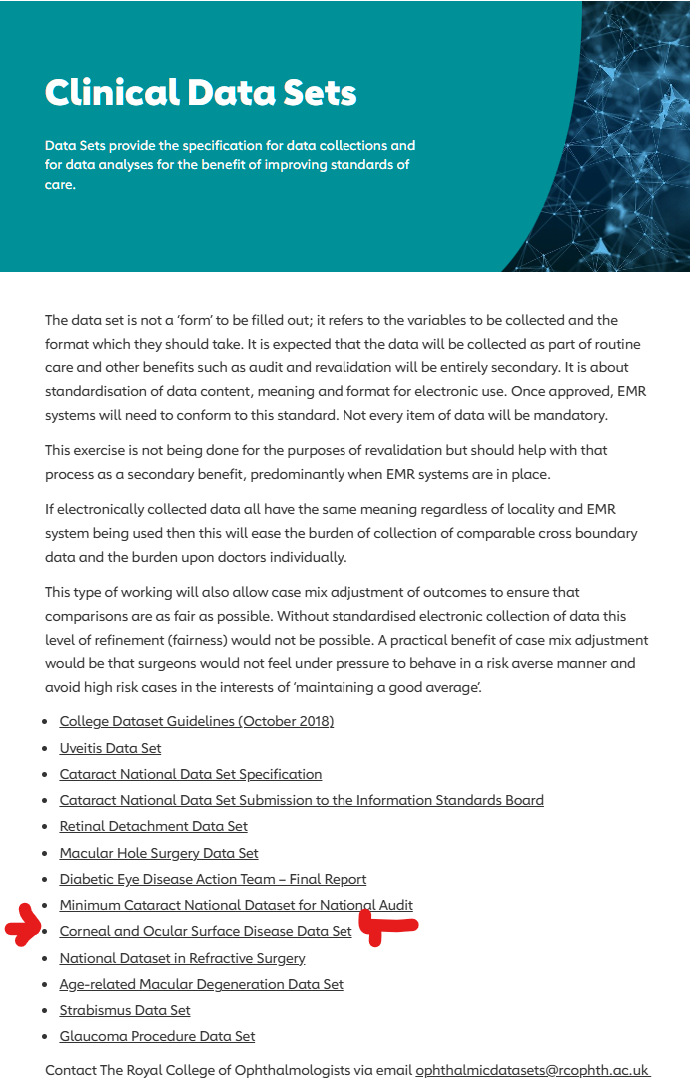

**Alternatively point your browser to:**

- <https://www.rcophth.ac.uk/standards-and-guidance/audit-and-data/clinical-data-sets/>
- Navigate to <<Corneal and Ocular Surface Disease Data Set>>
- Click to download file
